# Examining Migration, Social Bonds, Transnationalism, and HIV Prevention Pathways Among African Immigrants (MiST-Pathways): A Study Protocol

**DOI:** 10.64898/2026.05.27.26354266

**Authors:** Gloria Aidoo-Frimpong, Maame Araba Oduro, Portia Kamara, Donna Smith, the MiST-Pathways Study Team

**Affiliations:** Department of Epidemiology and Environmental Health, School of Public Health and Professions, University at Buffalo, State University of New York, Buffalo, NY, USA; Center for Interdisciplinary Research on AIDS, Yale University, New Haven, CT, USA; Transforming the HIV Response through Innovation and Equity (THRIVE) Lab at UB, Buffalo, New York, U.S.A; Department of Community Health, Ensign Global University, Akosombo, Ghana; Multicultural Community Family Services, Upper Darby, PA, USA

**Keywords:** HIV prevention, African immigrants, migration, relationships, HIV self-testing, pre-exposure prophylaxis, Palava Hut Conversations, mixed methods, community-engaged research, United States

## Abstract

**Background:** African immigrants in the United States bear a disproportionate HIV burden, with incidence approximately sixfold higher than the general population, yet remain largely absent from targeted prevention research. HIV vulnerability among this population is mediated through relationship and family systems that are restructured by migration, reorganizing household composition, gender norms, trust, and communication patterns through which prevention engagement occurs. Despite this, migration has rarely been examined as a force that transforms the relational contexts shaping engagement with HIV testing, HIV self-testing (HIVST), and pre-exposure prophylaxis (PrEP).

**Methods:** The MiST-Pathways Study will use a sequential mixed-methods, community-based pilot design among first-generation African immigrant adults (ages 18–50) residing in New York and Massachusetts. The study will proceed in three phases: Aim 1 will use semi-structured interviews (n = 15) and a structured survey (n = 75) to identify relationship typologies and migration-related relational mechanisms influencing HIV prevention engagement; Aim 2 will employ Palava Hut Conversations (PHC) (an African-centered deliberative method) with up to 30 participants to co-develop and prioritize relationship-tailored intervention components; and Aim 3 will conduct a proof-of-concept assessment of the prioritized component using a single-group pre–post design (n = 24), incorporating surveys and cognitive interviews to assess feasibility, acceptability, and preliminary evidence of mechanism activation. All activities will be conducted virtually via Zoom and WhatsApp, with eligibility screening administered through REDCap. The study has been approved by the University at Buffalo Institutional Review Board (STUDY00010347) and registered at ClinicalTrials.gov (NCT07565584).

**Discussion:** This protocol outlines the planned evaluation of a sequentially designed, community-engaged pilot study to examine how migration reshapes relational contexts that influence HIV prevention decision-making among African immigrants. Findings will inform the development of culturally grounded, relationship-tailored prevention strategies and the design of a future, larger-scale intervention study.

## Introduction

African immigrants in the United States (U.S.) bear a disproportionate HIV burden relative to their population size. Despite comprising less than 1% of the U.S. population, African immigrants experience HIV incidence approximately sixfold higher than the general population and nearly twofold higher than U.S.-born Black individuals [1]. African immigrants account for approximately 4% of national HIV diagnoses, with disproportionate representation among women and heterosexual transmission categories [2,3]. These disparities are amplified in Ending the HIV Epidemic (EHE) priority jurisdictions. In New York City, African immigrants account for approximately 14% of new HIV diagnoses; in Massachusetts, nearly 60% of HIV diagnoses among Black women occur among African immigrants [2,4]. Across these jurisdictions, African immigrants comprise up to 50% of HIV diagnoses among Black populations and up to 41% among women, placing this community at the epidemiologic center of the epidemic in several high-burden regions [5,6].

Despite this burden, African immigrants remain largely invisible in HIV surveillance and prevention programming due to routine aggregation within the broader Black or African American racial category [7]. This practice obscures population-specific epidemiology, exposure pathways, and prevention needs, limiting the precision of interventions in high-burden settings [2,7]. This invisibility has important implications for contemporary HIV prevention because African immigrants may not be adequately reached by interventions designed for the broader U.S. population or for aggregated Black communities. HIV self-testing (HIVST) and pre-exposure prophylaxis (PrEP) are central tools in contemporary HIV prevention [8–10]. HIVST can expand testing access by offering a private, flexible, and user-controlled option for individuals who may avoid clinic-based testing because of stigma, confidentiality concerns, or logistical barriers [11].

PrEP offers an effective biomedical prevention strategy for individuals at risk of HIV acquisition, including women and heterosexual adults who are often underrepresented in PrEP outreach [12]. For African immigrants, these approaches may be especially important because they create opportunities for prevention outside traditional clinical settings [9,10,13]. Yet their promise depends on whether they are visible, acceptable, discussable, and usable within the social and relational contexts in which prevention decisions occur.

Despite the promise of HIVST and PrEP, HIV prevention engagement among African immigrants remains shaped by intersecting individual, interpersonal, and structural factors [9,10]. At the individual level, inadequate HIV knowledge, low risk perception, and HIV-related stigma constrain prevention engagement [14,15]. At the interpersonal level, gender power imbalances, cultural norms around sexual communication, and partner dynamics limit women’s ability to negotiate safer sex and access prevention services [14,16]. Limited awareness and uptake of HIV self-testing (HIVST) and pre-exposure prophylaxis (PrEP) remain persistent challenges [9,10]. Community-level HIV stigma, structural barriers including language difficulties and health system navigation challenges, and gender-biased cultural norms compound risk and constrain prevention engagement [14–16]. These barriers do not operate in isolation. They are often filtered through relationships and family systems that shape how African immigrants understand HIV risk and engage with prevention[9]. Sexual partnerships, marriage, and family expectations influence what prevention options feel possible, acceptable, or safe to pursue [9,13]. Prevention decisions in this community are negotiated relationally, shaped by trust dynamics, communication norms, gender role expectations, and the perceived relational consequences of raising prevention within intimate relationships [9,17,18]. Research among East African immigrant women in the U.S. has documented that HIV testing carries profound relational stakes: requesting or disclosing a test can be interpreted by a partner as signaling infidelity or mistrust, transforming a routine clinical behavior into a relationship-threatening act [17]. Gender power inequalities further shape these dynamics; unequal power within partnerships creates differential safety around HIV disclosure, with women navigating competing risks between health and relational stability [18]. These findings point to a critical but under-addressed reality: in communities where relationships structure health decision-making, the relationship itself must become the unit of analysis for HIV prevention.

Migration reshapes these relationship systems and functions as a key social and structural determinant of health, transforming the relational contexts through which health decisions are made [19]. When individuals migrate, they undergo a reorganization of household composition, economic roles, gender norms, and decision-making authority [20]. Processes of separation, reunification, and prolonged transnational living alter the dynamics of trust, communication, and intimacy between partners [21]. Male labor migration disrupts the relational networks through which non-migrating partners navigate HIV prevention communication, altering support structures and the behavioral strategies available to those left behind [22]. Longitudinal population-based evidence shows that recent migration is associated with higher-risk sexual behaviors and increased HIV prevalence, with gender-differentiated mechanisms of vulnerability [23]. Among sub-Saharan African immigrants in European settings, HIV sexual risk behaviors occur within distinct relationship types, with reasons for risk-taking varying by relationship type and depending on cultural, social, and migration-related dimensions [24]. Migration does not directly produce HIV risk; rather, it restructures the relational context through which risk perception, disclosure, and prevention engagement occur [22–24]. Transnationalism, defined as the ongoing practices of communication, economic remittance, and identity maintenance that connect immigrants simultaneously to communities of origin and settlement [25] reinforces cultural norms governing HIV prevention within relationships. Transnational networks may transmit home-country stigma, norms of discretion around sexual health, and expectations about partnership roles across borders, shaping how African immigrants in the U.S. experience and respond to HIV prevention within their intimate relationships [14,16,25]. Yet no study has systematically examined how migration-driven relational restructuring, transnational social ties, and relationship typologies jointly shape HIV prevention pathways among African immigrants in the U.S.

We therefore propose the *MiST-Pathways Stud*y to address this critical gap. The study examines how migration-related experiences, social bonds, transnational connections, and intimate relationship dynamics shape HIV prevention pathways among African immigrants in the U.S. The *MiST-Pathways Study* is guided by a relational–socioecological framework that synthesizes two established theoretical traditions [26,27]. The socioecological model positions individual behavior within multiple levels of influence, individual, interpersonal, organizational, and structural, and has demonstrated that the relational level exerts among the strongest proximal influences on HIV prevention behavior among women in the U.S. [26,27]. Relationship science provides the conceptual foundation for understanding how relational processes such as trust, communication, power, and commitment function as operative mechanisms linking structural forces to individual prevention behavior [28]. Within this framework, migration-transformed relationships become the primary site of intervention, restructuring the relational conditions under which prevention decisions are made. Together, these perspectives allow the study to examine HIV prevention not only as individual health behavior, but as a relational and migration-shaped process embedded within families, partnerships, communities, and transnational networks.

## Materials and Methods

### Study Design

The MiST-Pathways Study is a sequential mixed-methods, community-based pilot study examining how migration-related relationship dynamics influence HIV prevention engagement among first-generation African immigrants. The study is guided by a relational–socioecological framework that positions relationships and how migration reshapes them as key mechanisms linking individual, interpersonal, and structural influences on HIV prevention behavior. The study will proceed in three sequential phases: Phase 1 identifies relationship typologies and relational mechanisms (Aim 1); Phase 2 co-develops and prioritizes relationship-tailored intervention components (Aim 2); and Phase 3 conducts a proof-of-concept assessment of the prioritized component (Aim 3). Each phase is directly informed by findings from the preceding phase.

### Study Status and Timeline

The study has not yet begun enrollment. IRB approval obtained in March 2026. Anticipated first enrollment is June 2026. The estimated study duration is 12 months. Months 1–3 will involve Aim 1 surveys and interviews; Months 4–6 will include Aim 2 Palava Hut Conversations; Months 7–10 will involve Aim 3 proof-of-concept procedures; and Months 11–12 will allow for data analysis and dissemination preparation. At the time of this protocol submission, recruitment and data collection have not yet begun, and no study outcomes have been generated.

### Ethical Considerations

This study has been reviewed and approved by the University at Buffalo Institutional Review Board (UB IRB; STUDY00010347), YALE Smart IRB (2000042465), and registered at ClinicalTrials.gov (NCT07565584). All study procedures will be conducted in accordance with the principles of the Declaration of Helsinki. The full IRB-approved study protocol is provided as Supporting Information (S2). Verbal informed consent will be obtained from all participants prior to enrollment. For the Aim 1 survey, participants will review and confirm an electronic consent statement programmed within REDCap before beginning. For all other activities, consent materials will be sent electronically via WhatsApp prior to each session, and verbal confirmation will be documented by study personnel. The UB IRB granted a waiver of written documentation of consent, consistent with the determination that the study presents no more than minimal risk and that participants are members of a distinct cultural group in which signing consent documents is not the norm. Participants will be informed of their right to withdraw at any time without penalty.

### Enrollment

The recommended schedule of enrollment, interventions, and assessments is presented in Fig 1 (See Supporting Information Figure 1). This includes enrollment procedures (screening and consent), intervention activities, and quantitative and qualitative assessments across all study timepoints.

A total of up to 144 participants will be enrolled across three aims: 90 in Aim 1 (75 survey, 15 interviews), 30 in Aim 2 (Palava Hut Conversations), and 24 in Aim 3 (proof-of-concept assessment). Approximately 175–200 individuals will be screened to achieve the target enrollment. As a pilot study, the trial is not powered for hypothesis testing; however, the qualitative sample of 15 interviews is consistent with established thematic saturation guidelines, and findings will inform the design and sample size calculations for a future, fully powered effectiveness study.

### Participants and Recruitment

#### Participants

Eligible participants must be first-generation African immigrants (born in an African country, currently residing in the U.S.), aged 18–50 years, residing in New York or Massachusetts, able to communicate in English, and have access to an internet-enabled device for Zoom and/or WhatsApp participation. Additional aim-specific criteria apply: Aim 2 participants must be willing to attend a 3–4-hour group deliberative session; Aim 3 participants must also match the prioritized relationship typology identified in Aim 2. Exclusion criteria include age under 18, non-African immigrant identity, residence outside New York or Massachusetts, inability to provide informed consent, and lack of required technology access. No exclusions will be made based on sex, gender identity, sexual orientation, income level, or immigration documentation status.

#### Recruitment

Recruitment will occur on a rolling basis during the first 2–4 months of the study through community-based organizations, faith-based organizations, civic and cultural associations, established African immigrant WhatsApp community groups, and electronic flyer distribution. IRB-approved electronic flyers containing study details and a secure REDCap screening link will be distributed by partner organizations and community leaders. Participants may also share flyers through word-of-mouth referral. The research team has previously demonstrated the feasibility of recruiting over 700 African immigrants within two months using similar WhatsApp-based strategies. All recruitment materials clearly state that participation is voluntary and research is conducted by researchers at the University at Buffalo.

#### Compensation

Participants will receive compensation proportional to their time commitment: $20 for Aim 1 survey completion; $40 for Aim 1 interview completion; $100 per Palava Hut Conversation session in Aim 2; $100 for Aim 3 full session completion; and an additional $40 for the optional cognitive interview in Aim 3. Compensation is not contingent upon completion of all study components.

### Study Procedures

#### Aim 1 - Identification of Relationship Typologies and Mechanisms

Two data collection activities will occur. First, 75 participants will complete a brief structured survey (15–20 minutes) administered electronically via REDCap, collecting data on demographics, relationship characteristics, migration experiences, and HIV testing, HIVST, and PrEP awareness and behaviors. Survey findings will be summarized descriptively to triangulate qualitative data. Second, 15 participants will complete a semi-structured interview via Zoom or WhatsApp (60–90 minutes) examining relationship structure, communication norms, HIV prevention decision-making, and migration experiences including separation, reunification, and transnational ties. Interviews will be audio-recorded, transcribed, and de-identified. Recruitment and survey administration will occur concurrently, with thematic analysis beginning immediately after interview completion to inform Aim 2.

#### Aim 2 - Palava Hut Conversations for Co-Development and Prioritization

Three Palava Hut Conversations (PHCs) will be conducted after preliminary Aim 1 analysis. PHC is an African-centered deliberative method through which participants gather to share experiences, deliberate collectively, and make decisions in a guided environment. PHC 1 (New York, n = 10) and PHC 2 (Massachusetts, n = 10) will each involve facilitated discussion on relationship dynamics, HIV prevention decision-making, migration experiences, and trust and communication norms, followed by presentation of Aim 1 findings for collective validation of typologies and generation of candidate intervention components (3–4 hours each, via Zoom). PHC 3 (n = 20–30, same participants reconvened) will synthesize outputs from PHC 1 and 2. Participants will engage in silent reflection before discussion, then use a structured prioritization matrix to rate candidate components on cultural congruence, feasibility, and impact using a 1–5 scale. Scores will be recorded in real time by the facilitator, and the highest-ranked component will advance to Aim 3. **Aim 3 - Proof-of-Concept Assessment:** Participants matching the prioritized relationship typology (n = 24) will complete a single structured session (3–5 hours) via Zoom and REDCap consisting of four sequential steps: (1) baseline survey (T0, ∼10–12 minutes) assessing HIV testing intention, HIVST willingness, PrEP interest, and relationship communication readiness; (2) standardized intervention exposure session (∼60–90 minutes) delivering the component prioritized in Aim 2 through structured communication prompts, scenario-based discussion, or guided reflection exercises; (3) post-intervention survey (T1, ∼15–20 minutes) assessing the same constructs plus perceived intervention relevance; and (4) optional cognitive interview (∼60–75 minutes) for a self-selected subset, assessing acceptability, clarity, feasibility, and mechanism activation.

Table 1 summarizes the data collection activities, methods, and measures across all three study aims.

**Table 1.**
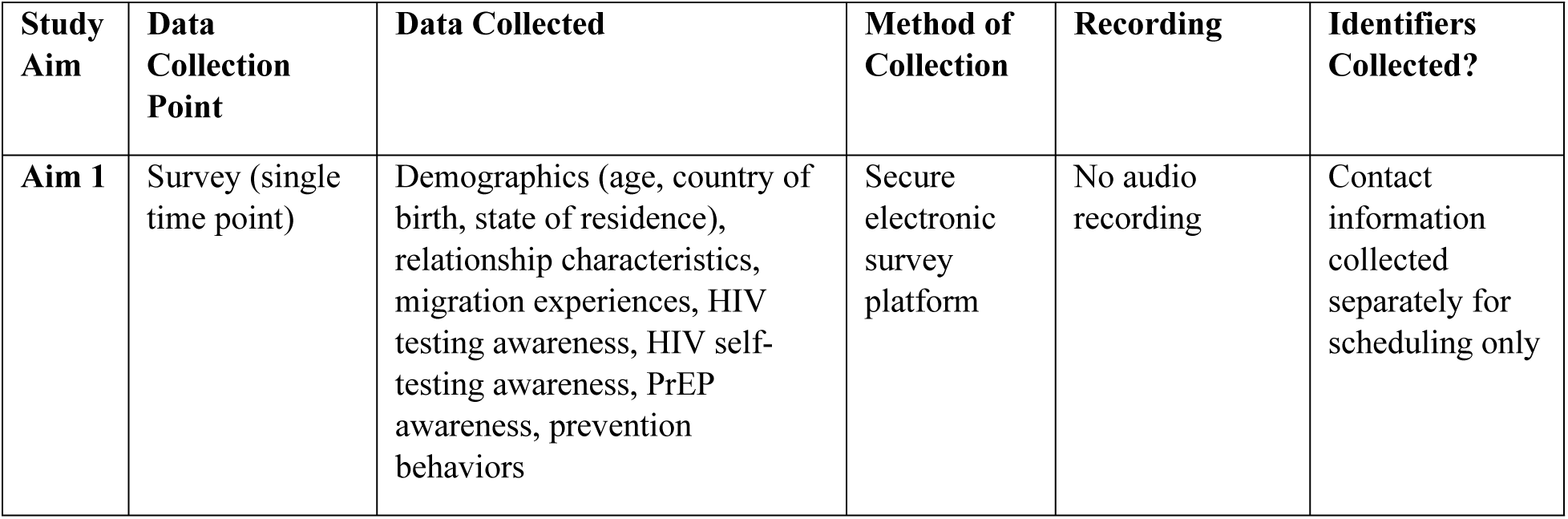
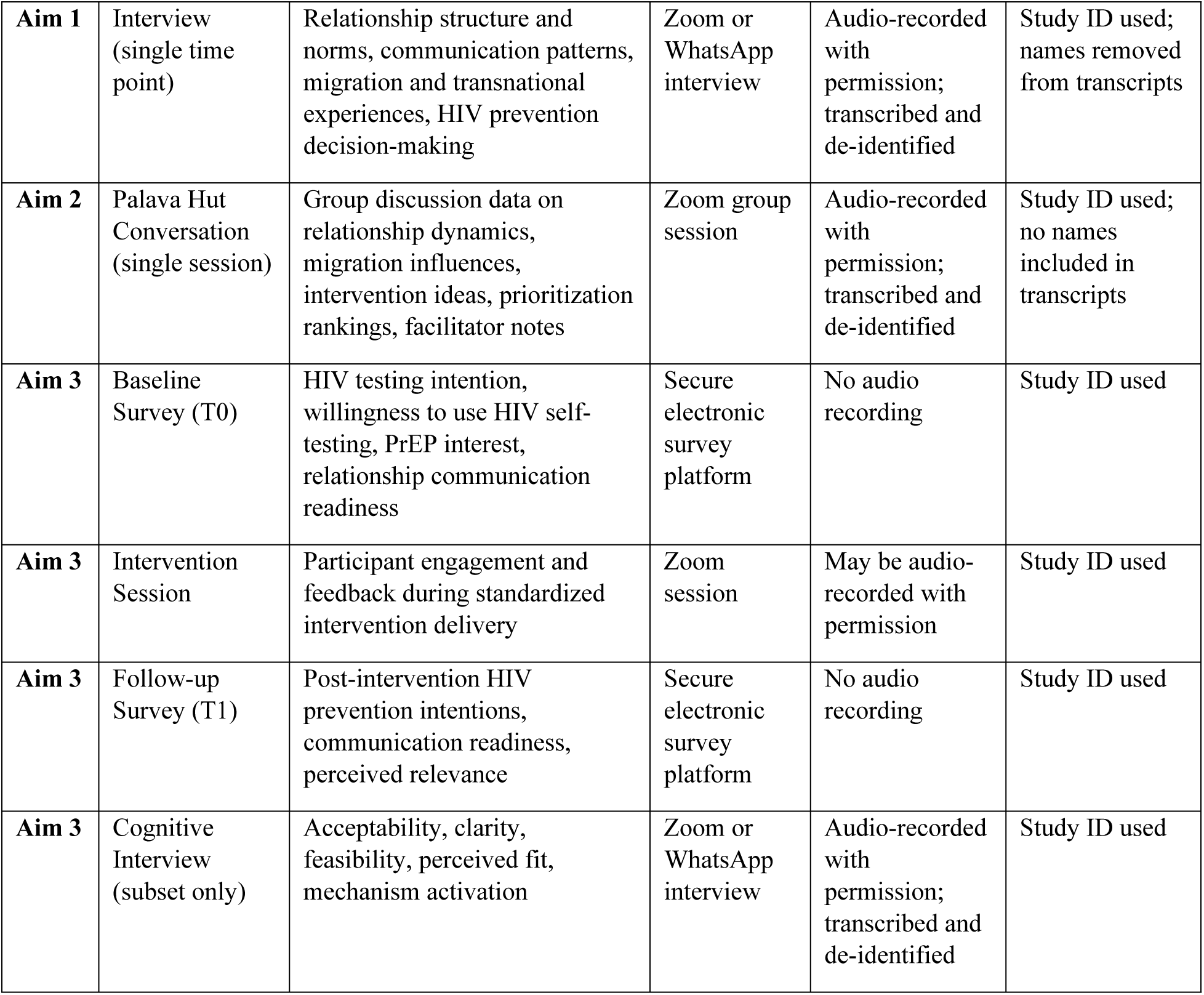
Data collection activities, methods, and measures by study aim.

### Data Collection and Analysis

#### Quantitative Data

Survey data will be collected via REDCap at each time point. Aim 1 survey data and Aim 3 baseline (T0) and post-intervention (T1) data will be summarized using descriptive statistics. Aim 3 analysis will assess the direction and consistency of pre–post change in HIV testing intention, HIVST willingness, PrEP interest, and relationship communication readiness. The proof-of-concept assessment is focused on the feasibility, acceptability, and plausibility of mechanism activation rather than hypothesis testing. Acceptability, feasibility, and appropriateness will be assessed using validated implementation outcome measures, each consisting of a four-item scale administered at T1 [29]. An additional measure of acceptability will be a pre-specified criterion, where the intervention will be considered acceptable if 75% or more of participants rate it favorably.

### Qualitative Data

Aim 1 interview transcripts will be analyzed using rapid qualitative analysis with rapid synthesis matrices to identify relationship typologies and migration-related relational mechanisms. Aim 2 analysis will occur primarily in real time through facilitated deliberation, with audio recordings, facilitator notes, decision logs, and prioritization matrices synthesized using rapid qualitative approaches to validate typologies, identify modifiable mechanisms, and rank intervention components. Aim 3 cognitive interview data will undergo rapid thematic analysis focused on acceptability, feasibility, appropriateness, perceived fit, usability, and evidence of mechanism activation.

### Mixed-Methods Integration

Findings will be integrated sequentially across aims. Aim 1 findings will inform Aim 2 deliberation and component prioritization. The prioritized component will then be evaluated in Aim 3, with qualitative and quantitative findings triangulated to inform refinement and future study design. Quality control procedures will include pilot testing of survey instruments, review of data for completeness, and verification of transcripts against audio recordings.

### Data Management Plan

All study data will be collected and stored on secure University at Buffalo systems. Eligibility screening and survey data will be collected and stored in REDCap, a secure web-based application hosted by the University at Buffalo on institutionally managed servers. Audio recordings, deidentified transcripts, and analytic datasets will be stored on a secure, password-protected University at Buffalo network drive. Study documents may also be stored on University at Buffalo–approved cloud platforms, including UB Box and UB OneDrive, which require university authentication and role-based access controls. No study data will be stored on personal devices, unencrypted external drives, or non–University at Buffalo–approved cloud storage systems.

Each participant will be assigned a unique study identification number. Identifiable information, including names, email addresses, and telephone numbers, will be stored separately from research data in a restricted-access file. A code key linking participant identifiers to study identification numbers will be maintained in a separate, password-protected file accessible only to the Principal Investigator and authorized study personnel. The code key will be destroyed once data analysis is complete and there is no longer a need to re-contact participants, or within three years after study closure, whichever occurs first. De-identified electronic research records will be retained for a minimum of three years after study closure in accordance with federal regulations and may be retained longer for future secondary analyses or grant development, consistent with institutional policy.

### Expected Outcomes

The MiST-Pathways Study is expected to generate foundational, mechanism-focused evidence on how migration-related relational dynamics shape engagement with HIV testing, HIVST, and PrEP among African immigrants. Table 2 summarizes the expected outcomes across all three study aims. *Table 2. Expected outcomes by study aim*

**Table 2.** Expected outcomes by study aim

| Outcome Category | Outcome | Measure | Time Frame |
| --- | --- | --- | --- |
| **Primary Outcomes**<br>**(Aim 3)** | Acceptability of Intervention | Mean AIM score (1–5 scale);<br>75% acceptability threshold | T1 - immediately post-intervention |
|  | Feasibility of Intervention | Mean FIM score (1–5 scale) | T1 - immediately post-intervention |
|  | Appropriateness of Intervention | Mean IAM score (1–5 scale) | T1 - immediately post-intervention |
| **Secondary Outcomes (Aim 3)** | Change in HIV Testing Intention | Mean change score (1–5 scale) | T0 to T1 |
|  | Change in HIV Self-Testing Willingness | Mean change score (1–5 scale) | T0 to T1 |
|  | Change in Communication Readiness | Mean change score across 3 items (1–5 scale) | T0 to T1 |
|  | Change in PrEP Awareness and Interest | Mean change score (1–5 scale) | T0 to T1 |
| **Other Pre-specified Outcomes (Aim 1)** | Relationship Typologies | Number of typologies identified; reported narratively | Months 1–3 |
|  | HIV Prevention Awareness and Behaviors | Frequencies and proportions (%) | Months 1–3 |

## Discussion

The MiST-Pathways Study protocol addresses a critical gap in HIV prevention research by generating foundational, mechanism-focused evidence on how migration-related relational dynamics shape engagement with HIV testing, HIVST, and PrEP among first-generation African immigrants in the U.S. Despite the disproportionate HIV burden borne by this population, African immigrants remain largely absent from targeted prevention research, and the relational pathways through which migration restructures HIV prevention decision-making have not been systematically characterized [2,7]. This study directly responds to that gap by integrating rigorous qualitative inquiry, African-centered community deliberation, and a proof-of-concept behavioral assessment within a single sequential mixed-methods design.

The significance of this study lies in two interconnected theoretical and public health arguments that remain underdeveloped in existing HIV prevention science. First, migration is an active, ongoing social force that restructures the relational conditions under which HIV prevention decisions are made [19,20]. For African immigrants, migration reorganizes household composition, gender roles, and decision-making authority. This reorganization introduces separation, reunification, and transnational living as distinct relational stressors that alter the dynamics of intimacy, trust, and communication between partners. These disruptions also embed individuals within cross-border networks that reinforce cultural norms of discretion, respectability and stigma around HIV [19–21]. Collectively, these migration-driven relational shifts fundamentally shape whether and how individuals engage with HIV testing, HIVST, and PrEP [17,18,22,23]. While HIV prevention frameworks have addressed individual and structural factors, sustained attention to the relational level, the partnership dynamics, communication norms, and trust that mediate between structural context and individual behavior, has been limited, particularly among migrant populations [18,22,24,26]. This study positions that relational level as the primary intervention target. Relationship typologies matter here because not all relationships carry the same HIV prevention implications. Migration, transnational obligation, and cultural renegotiation reshape relational contexts in ways that produce real variation in how people engage with testing and prevention [19,21,25]. For African immigrants, characterizing that variation is the foundation for prevention strategies that reflect how people actually live and make health decisions after migration.

Several limitations of this study are worth noting, along with how the team has planned around them. The study is limited to New York and Massachusetts, which means findings may not transfer directly to African immigrant communities in other parts of the country. Both states are among the highest-burden EHE jurisdictions for this population and include participants from diverse countries of origin and migration backgrounds. Participation is virtual, which could exclude people with limited internet access or digital comfort. However, prior work has demonstrated the feasibility of WhatsApp and Zoom within this community, and technical support will be available throughout [30]. Another limitation is the inclusion of only English-speaking participants, which may miss more recently arrived immigrants. The study requires substantive verbal participation in group deliberation, making this restriction a practical necessity. However, most eligible African immigrants have at least basic English proficiency, thereby reducing this barrier and broadening the study’s reach. Future work will pursue multilingual adaptations to include those not yet reached. As a pilot study, the study is not powered for hypothesis testing. Rather, the purpose of this study is to establish feasibility, acceptability, and appropriateness to generate directional evidence. This will help produce community-grounded data needed to design a definitive intervention.

### Dissemination

Findings will be disseminated through a structured, community-centered strategy prioritizing direct engagement with African immigrant communities through WhatsApp-ready video messages, a digital flip book, and a virtual community dialogue session (anticipated n = 40–60), all co-developed with MCFS. Findings will also be shared through peer-reviewed journal articles, conference presentations, and community feedback sessions with participating organizations and networks. Dissemination materials will be hosted on the African Immigrant Health Research Collaborative (AIHRC) website and distributed through established WhatsApp networks. Due to the sensitive nature of the data, individual participant data will not be shared publicly; all data will be de-identified and stored securely on password-protected University at Buffalo systems accessible only to the research team.

### Amendments and Study Termination

Any modifications to the study protocol, including changes to eligibility criteria, study procedures, data collection instruments, or consent documents, will be submitted to the University at Buffalo IRB for review and approval before implementation. Modifications will be documented and tracked through the IRB submission system. In the event that the study must be terminated early, due to safety concerns, funding discontinuation, or other unforeseen circumstances, enrolled participants will be notified, all data collection will cease, and data already collected will be retained and secured in accordance with the University at Buffalo data management policies and federal regulations. The IRB will be notified of any early termination in accordance with applicable institutional requirements.

## Data Availability

No datasets were generated or analyzed during the current study because this manuscript describes a study protocol. De-identified research data generated from the pilot implementation study described in this protocol will be made publicly available upon study completion and publication of the primary findings, in accordance with institutional review board approvals and participant consent procedures.

## Author Contributions

**Conceptualization:** Gloria Aidoo-Frimpong, Portia Kamara, Donna Smith

**Funding Acquisition:** Gloria Aidoo-Frimpong

**Methodology:** Gloria Aidoo-Frimpong, Portia Kamara, Donna Smith

**Writing- original draft:** Gloria Aidoo-Frimpong, Maame Araba Oduro

**Writing- review and editing:** Gloria Aidoo-Frimpong, Maame Araba Oduro, Portia Kamara, Donna Smith

## Acknowledgments

The authors wish to thank the African immigrant community organizations and networks in New York and Massachusetts, whose continued partnership makes this research possible. We are grateful to participants who will contribute their time and experiences to this work.

## Funding

Research reported in this publication was supported by award number P30MH062294 from the National Institute of Mental Health to the Yale University Center for Interdisciplinary Research on AIDS. The content is solely the responsibility of the authors and does not necessarily represent the official views of the Center for Interdisciplinary Research on AIDS, the National Institute of Mental Health, or the National Institutes of Health.

## Competing Interest

The authors have no competing interests.

## Supporting Information

S1 File. SPIRIT 2025 checklist

S1File. Figure 1 SPIRIT Study schedule of enrollment, interventions, and assessments S2 File. Detailed protocol submitted to the ethics committee

## Notes

### Competing Interest Statement

The authors have declared no competing interest.

### Clinical Trial

NCT07565584

### Funding Statement

Yes

### Author Declarations

This study has been reviewed and approved by the University at Buffalo Institutional Review Board (UB IRB STUDY00010347), YALE Smart IRB (2000042465), and registered at ClinicalTrials.gov (NCT07565584).

